# Machine learning enabled subgroup analysis with real-world data to inform better clinical trial design

**DOI:** 10.1101/2021.05.11.21257024

**Authors:** Jie Xu, Hao Zhang, Hansi Zhang, Jiang Bian, Fei Wang

## Abstract

Restrictive eligibility criteria for clinical trials may limit the generalizability of treatment effectiveness and safety to real-world patients. In this paper, we propose a machine learning approach to derive patient subgroups from real-world data (RWD), such that the patients within the same subgroup share similar clinical characteristics and safety outcomes. The effectiveness of our approach was validated on two existing clinical trials with the electronic health records (EHRs) from a large clinical research network. One is the donepezil trial for Alzheimer’s disease (AD), and the other is the Bevacizumab trial on colon cancer (CRC). The results show that our proposed algorithm can identify patient subgroups with coherent clinical manifestations and similar risk levels of encountering severe adverse events (SAEs). We further exemplify that potential rules for describing the patient subgroups with less SAEs can be derived to inform the design of clinical trial eligibility criteria.

## Introduction

Randomized controlled trials (RCTs) are the golden standard for investigating drug effectiveness (1). In designing an RCT, stringent eligibility criteria (EC) need to be applied to appropriately define a study population so that the drug effectiveness can be reliably and safely evaluated. In practice, trial designers often adopt eligibility criteria from existing similar trials, without too much consideration on their applicability and adaptability to the new trials. Moreover, many pivotal Phase III clinical trials continue to apply the same highly restrictive eligibility criteria as in the corresponding Phase I and Phase II trials. These oftentimes make the eligible trial participants not representative of the real-world patient population who will receive the treatment (2). For example, the elderlies are often excluded from cancer and Alzheimer’s disease drug development trials and are therefore under-represented among the primary target populations for these drugs (3, 4).

Although excessive or overly restrictive EC may lower the risk of the study populations for encountering adverse events (5-7), they usually lead to low population representativeness (thus, low trial generalizability), and subsequently, treatment effectiveness could be reduced, and the likelihood of adverse outcomes could increase when the treatment entered real-world clinical practice (8). Therefore, as recommended by regulatory agencies such as the U.S. Food and Drug Administration (FDA), broadening eligibility criteria during enrollment to increase the diversity of the clinical trial population and thus improve generalizability is an important trend (9).

Trial generalizability is largely dependent on the representativeness of the study population with respect to target population to which the study results are intended to be applied (8). In recent years, the rapid adoption of electronic health records (EHR) systems has led to large integrated clinical data warehouses and interoperable data networks, which made it possible for accumulating large amounts of real-world patient data. Such data provide us a unique opportunity for simulating the study and target populations of clinical trials. The goal of this study is to develop machine learning approaches for mining insights from RWD that could be informative of clinical trial eligibility criteria design. In particular, to account for the heterogeneity of the real-world population, we introduce a novel transparent and outcome-guided probabilistic model to identify the subphenotypes who took a specific medication (Fig. 1). The patients within the same subphenotype do not just share similar clinical characteristics, but also face a similar risk of encountering severe adverse events (SAEs) after they take the drug. We assume certain compositions (co-occurrence patterns) of the clinical events within an individual’s EHR can determine those subphenotypes and propose a novel weakly supervised clinical topic modeling approach to identify those subphenotypes. Here each clinical topic represents a certain clinical event composition pattern learned from the patient EHRs.

**Fig. 1.**
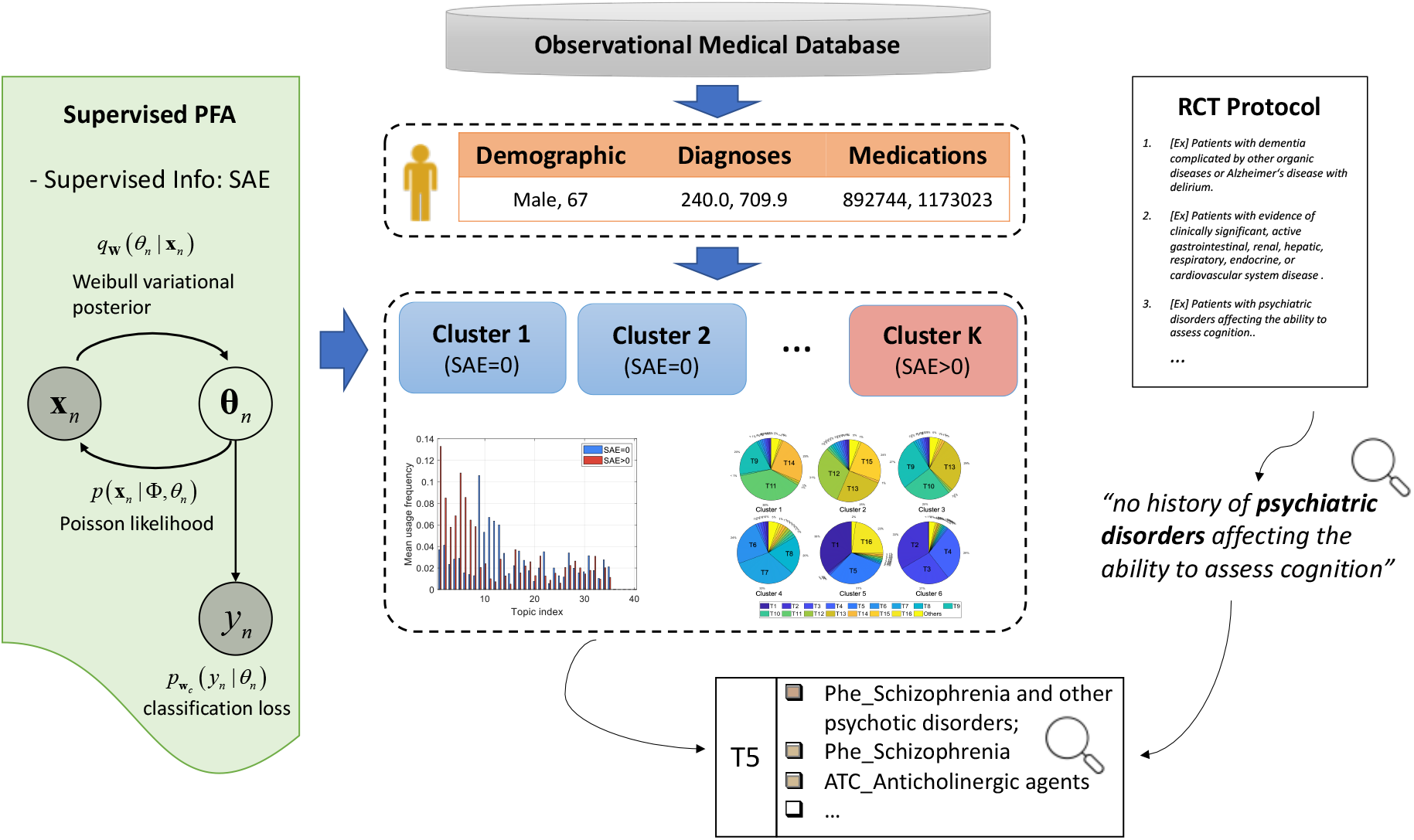
Model overview. Clinical events including demographics, diagnoses, and medications were extracted from RWD to represent patients. Supervised Poisson factor analysis (PFA) is applied to identify patient subgroups with coherent clinical latent topics and outcomes measured by SAEs. Subgroups with less SAEs can be derived to inform the design of clinical trial eligibility criteria.

We evaluated our algorithm with two real-world clinical trials: (1) a hallmark randomized Phase III trial (i.e., NCT00478205) that compares the effects of 23 mg to 10 mg Donepezil in treating patients with Alzheimer’s disease that led to the FDA approval of the 23 mg Donepezil; and (2) another randomized Phase III trial studying two different combination chemotherapy regimens with or without bevacizumab (trade name Avastin) in high-risk stage II/III colon cancer patients. We used RWD data from the OneFlorida Clinical Research Consortium -- a statewide clinical data warehouse containing large collections of linked EHR, administrative claims, vital statistics, and cancer registry data among others, covering more than 15 million (∼60%) Floridians (10). Our results showed that, for both trials, 1) a significant number of real-world patients who satisfied the ECs and took the drug suffered from SAEs; 2) our model can clearly identify the patient subgroups who are more likely to suffer or not suffer from SAEs as subphenotypes in a transparent and interpretable way. We have also inferred the clinical topics from the subphenotypes with or without SAEs, which reveal meaningful combinations of clinical features across multiple data types and provide data-driven recommendations for refining the ECs of clinical trials.

## Results

### Target population of the donepezil clinical trial

NCT00478205 is a double-blind, double-dummy trial that compares different dosages (23 mg vs. 10 mg) of donepezil for treating patients with moderate to severe Alzheimer’s disease (11). The real-world individual-level EHR data are from the OneFlorida Clinical Research Consortium. We constrained the target population as those who (1) were diagnosed with AD, and (2) treated with donepezil. A total of 4,998 unique patients (mean (SD) age, 77.53 (9.9) years) were identified from OneFlorida (Fig. 2b, Table 1).

**Table 1.** Demographic characteristics and selected traits of the target population of the donepezil trial for Alzheimer’s disease.

| Characteristic | Overall<br>(N=4,998) | # SAEs = 0<br>(N=3,063) | # SAEs > 0<br>(N=1,935) | $\chi^2$<br>p-value | Adjust Demo* |
| --- | --- | --- | --- | --- | --- |
| Age, Mean (SD), yr | 77.53 (9.9) | 76.98 (9.8) | 78.41 (9.9) |  |  |
| Sex, No. (%) |  |  |  |  |  |
| Female | 3,123 (62.5) | 1,923 (62.8) | 1,200 (62.0) |  |  |
| Race/Ethnicity, No. (%) |  |  |  |  |  |
| White | 3,537 (70.8) | 2,262 (73.8) | 1,275 (65.8) |  |  |
| Black | 965 (19.4) | 484 (158) | 481 (24.9) |  |  |
| Asian | 34 (0.6) | 25 (0.8) | 9 (0.5) |  |  |
| Others & Unknown | 462 (9.2) | 292 (9.5) | 170 (8.8) |  |  |
| Study traits, No. (%) |  |  |  |  |  |
| Memantine | 1,511 (30.2) | 950 (31) | 561 (28.9) | 0.129 | 0.145 |
| Psychiatric disorders | 1,396 (27.9) | 754 (24.6) | 642 (33.1) | $\leq 0.001$ | $\leq 0.001$ |
| Cardiovascular (CS*) | 1,082 (21.6) | 492 (16) | 590 (30.4) | $\leq 0.001$ | $\leq 0.001$ |
| Endocrine (CS*) | 813 (16.2) | 353 (11.5) | 460 (23.7) | $\leq 0.001$ | $\leq 0.001$ |
| Cancer | 808 (16.1) | 468 (15.2) | 340 (17.5) | 0.032 | 0.091 |
| Dysphagia | 649 (12.9) | 302 (9.8) | 347 (17.9) | $\leq 0.001$ | $\leq 0.001$ |
| Gastrointestinal (CS*) | 631 (12.6) | 289 (9.4) | 342 (17.6) | $\leq 0.001$ | $\leq 0.001$ |
| Drug or alcohol abuse<br>or dependence | 627 (12.5) | 283 (9.2) | 344 (17.7) | $\leq 0.001$ | $\leq 0.001$ |
| Respiratory (CS*) | 586 (11.7) | 249 (8.1) | 337 (17.4) | $\leq 0.001$ | $\leq 0.001$ |
| AD with delirium | 389 (7.7) | 161 (5.2) | 228 (11.7) | $\leq 0.001$ | $\leq 0.001$ |
| Hepatic disease | 361 (7.2) | 180 (5.8) | 181 (9.3) | $\leq 0.001$ | $\leq 0.001$ |
| Renal (CS*) | 342 (6.8) | 135 (4.4) | 207 (10.6) | $\leq 0.001$ | $\leq 0.001$ |
| Parkinson disease | 329 (6.5) | 176 (5.7) | 153 (7.9) | 0.003 | 0.001 |
| Menopausal | 230 (4.6) | 128 (4.1) | 102 (5.2) | 0.073 | 0.040 |
| Antidepressant | 226 (4.5) | 143 (4.6) | 83 (4.2) | 0.530 | 0.590 |
| Basal/squamous<br>cell carcinoma of the skin | 216 (4.3) | 128 (4.1) | 88 (4.5) | 0.532 | 0.275 |
| Gastric ulcers | 163 (3.2) | 75 (2.4) | 88 (4.5) | $\leq 0.001$ | $\leq 0.001$ |
| Inflammatory bowel disease | 154 (3) | 82 (2.6) | 72 (3.7) | 0.038 | 0.024 |
| Rivastigmine | 153 (3) | 121 (3.9) | 32 (1.6) | $\leq 0.001$ | $\leq 0.001$ |
| Multi-infarct dementia | 151 (3) | 72 (2.3) | 79 (4) | $\leq 0.001$ | 0.001 |
| Acupressure | 119 (2.3) | 54 (1.7) | 65 (3.3) | $\leq 0.001$ | 0.002 |
| Fecal incontinence | 107 (2.1) | 48 (1.5) | 59 (3) | $\leq 0.001$ | 0.001 |
| Galantamine | 35 (0.7) | 18 (0.5) | 17 (0.8) | 0.230 | 0.190 |
| Severe lactose intolerance | 24 (0.4) | 12 (0.3) | 12 (0.6) | 0.255 | 0.219 |
| Hepatic (CS*) | 20 (0.4) | 9 (0.2) | 11 (0.5) | 0.134 | 0.105 |

CS\*: Clinically significant. If the disease causes hospitalization, we consider it as "clinically significant". Demo\*: Demographics (i.e., age, gender and race).

**Fig. 2.**
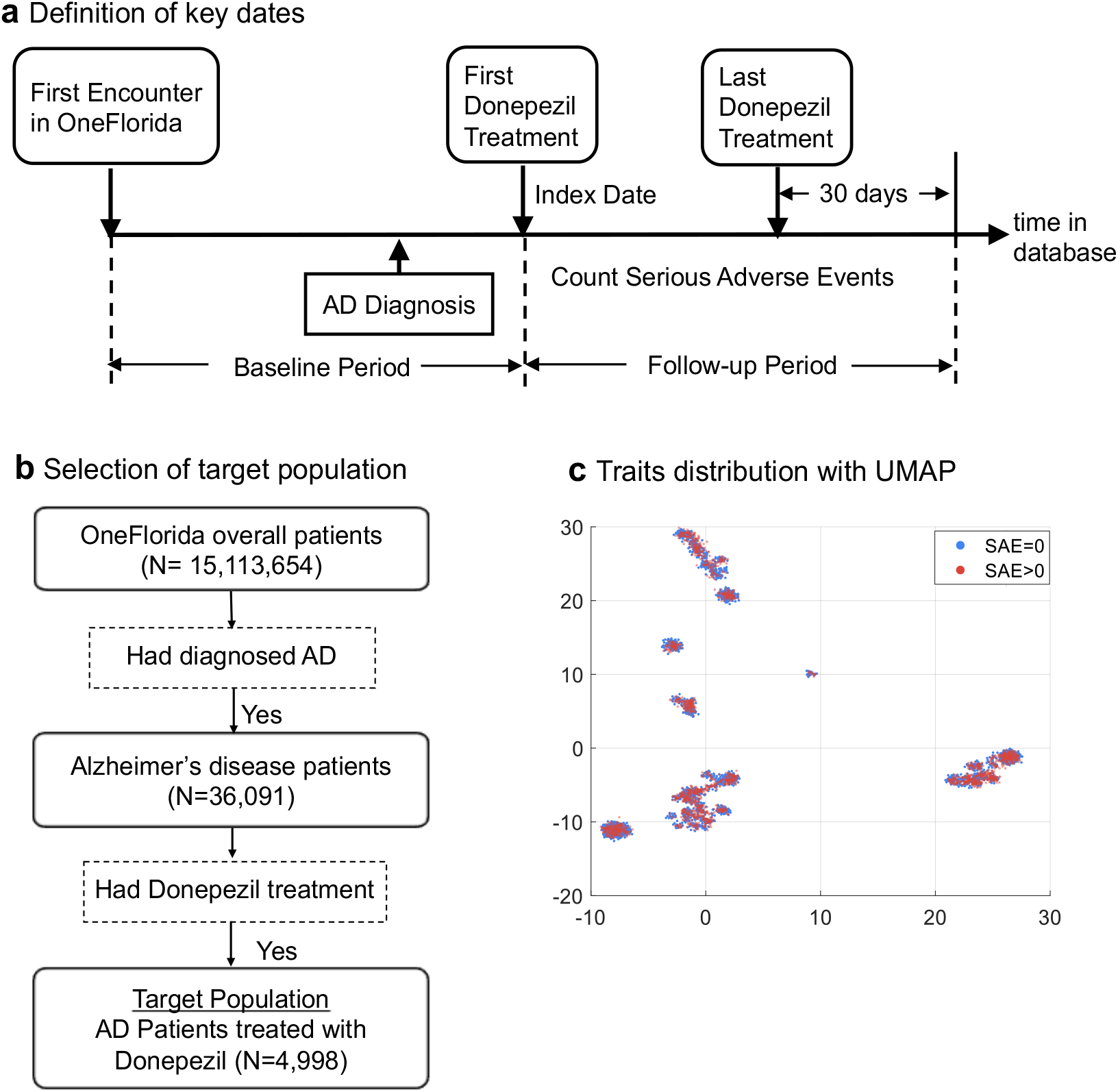
The donepezil trial for Alzheimer’s disease. **a** Definition of key dates; **b** Selection of target population; **c** Traits distribution with UMAP among two patients’ groups with or without SAE.

To determine whether a patient experienced SAEs related to donepezil, we first compiled the list of SAEs reported in the original trial in ClinicalTrials.gov, and then identified relevant diagnosis codes (i.e., International Classification of Diseases, Ninth/Tenth Revision, Clinical Modification [ICD-9/10-CM]) for each SAE. Based on the severity grading scale defined in the Common Terminology Criteria for Adverse Events (CTCAE) (12), adverse events leading to hospitalization or mortality are grade 3 and above are considered as SAEs in our study. Further, we defined that the SAE diagnosis codes have to occur 1) after the first donepezil treatment, and 2) before 30 days after the last donepezil treatment (Fig. 2a). Among the patients in our target population, 3,063 (61.3%) had no SAE while 1,935 (38.7%) had at least one SAE. We dropped the ECs which are not computable (e.g., subjective eligibility criteria such as “written informed consent” is not computable, but also not going to affect our analysis, since we can safely assume all patients treated with donepezil in the real-world are consented) and extracted study traits corresponding to each computable EC from the OneFlorida data (Table 1).

We first represent each patient using the extracted traits as a vector and check whether patients with and without SAEs can be well separated. Fig. 2c shows the 2D embeddings of patient traits with Uniform Manifold Approximation and Projection (UMAP) (13). We color each sample based on whether the patient had SAEs or not. As shown in the figure, the two clusters, patients with (#SAE>0) vs. without (#SAE=0) are mixed up with each other, which indicates that the trial ECs cannot guarantee the safety for real-world patients. In addition, we examined the differences of the study traits value distributions between the two groups through Chi-square tests and summarized the results in Table 1, from which we observe that many traits were not significantly different (statistical sense, with p>0.05) including memantine (p=0.145), antidepressant (p=0.590), basal/squamous cell carcinoma of the skin (p=0.275), galantamine (p=0.190), severe lactose intolerance (p=0.219), and clinically significant Hepatic (p=0.105). This suggests that the trial ECs can further be optimized with real-world evidence.

### Study design of the donepezil clinical trial

The beginning of the treatment is termed the index date. We set the index date in our donepezil trial to the first (ever) observed prescription date of donepezil. We refer to the observed time before the index date as the baseline period and use the information (i.e., demographic, diagnosis and medication history) collected during that time period for analysis. The period from the index date to the last donepezil prescription plus 30 days was set as the follow-up period, from which the SAE information is collected. Fig. 2a illustrates the key dates in our study design.

We applied supervised Poisson factor analysis (SPFA) to the collected information and used the occurrence of SAE as the supervision to guide the learning process of PFA. Similar to other topic modeling approaches (14), SPFA first compressed the clinical events into a set of overlapping groups (i.e., topics). Novel patient representations are derived on top of these topics (which are groups of clinical events that tend to co-appear in the same visit within the RWD). K-means clustering is then performed on these new patient representations to identify the clusters as subphenotypes. To choose the optimal number of topics K, we used all samples to learn the supervised topic model and then evaluated the topic coherence by normalized pointwise mutual information (NPMI) value (15), and the classification performance by ROC-AUC. We set K=40 for subsequent analyses as it achieved the highest ROC-AUC with large topic coherence values (Supplementary Fig. 1). The model robustness with respect to K will be discussed in the methods.

As discussed in (16), according to the silhouette score (17), we select the most appropriate number of clusters that provides the largest silhouette score. As a result, six clusters were derived with K-means for patients who had been diagnosed with AD and treated with Donepezil (Fig. 3a). Among the six clusters, two patient subgroups emerged: (1) the SAE group (#SAE>0) containing clusters 4, 5, and 6, and (2) the non-SAE group (#SAE=0) including clusters 1, 2 and 3. As shown in Fig. 4a, the two patient subgroups (i.e., #SAE=0 vs. #SAE>0) are well separated. Specifically, for the SAE group, 1,915 out of total 1,935 patients (99.0%) encountered SAE events; for non-SAE group, 3,014 out of 3,063 patients (98.4%) did not have any SAE events.

**Fig. 3.**
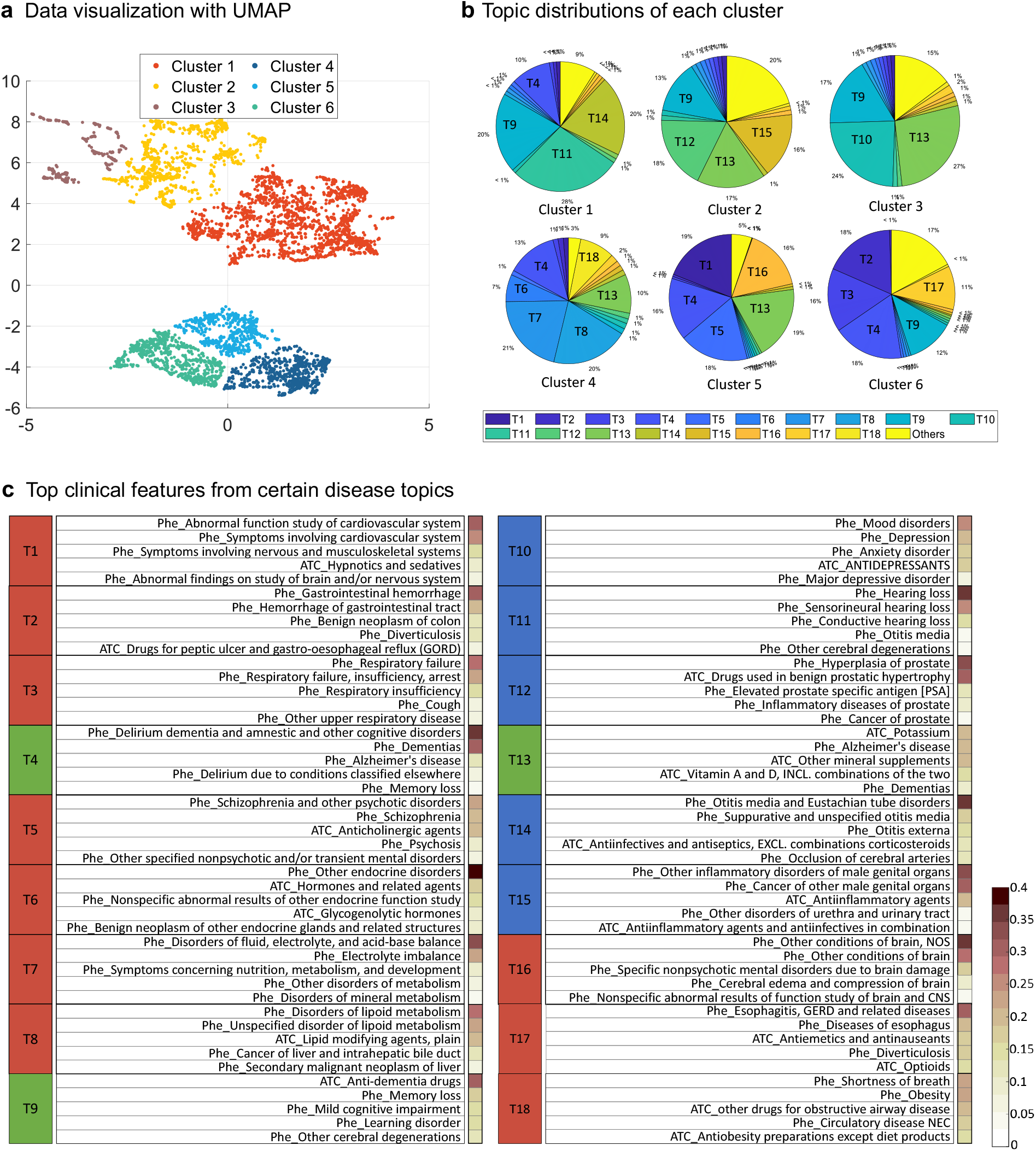
Clustering results of AD target population. **a** Data visualization with UMAP; **b** Topic distributions of each cluster; **c** Top clinical features from certain disease topics, where the red, blue, and green topics represent the commonly used topics for clusters 4, 5, 6 (most of samples are with SAE>0), clusters 1, 2, 3 (most of samples are with SAE=0), and all clusters, respectively.

**Fig. 4.**
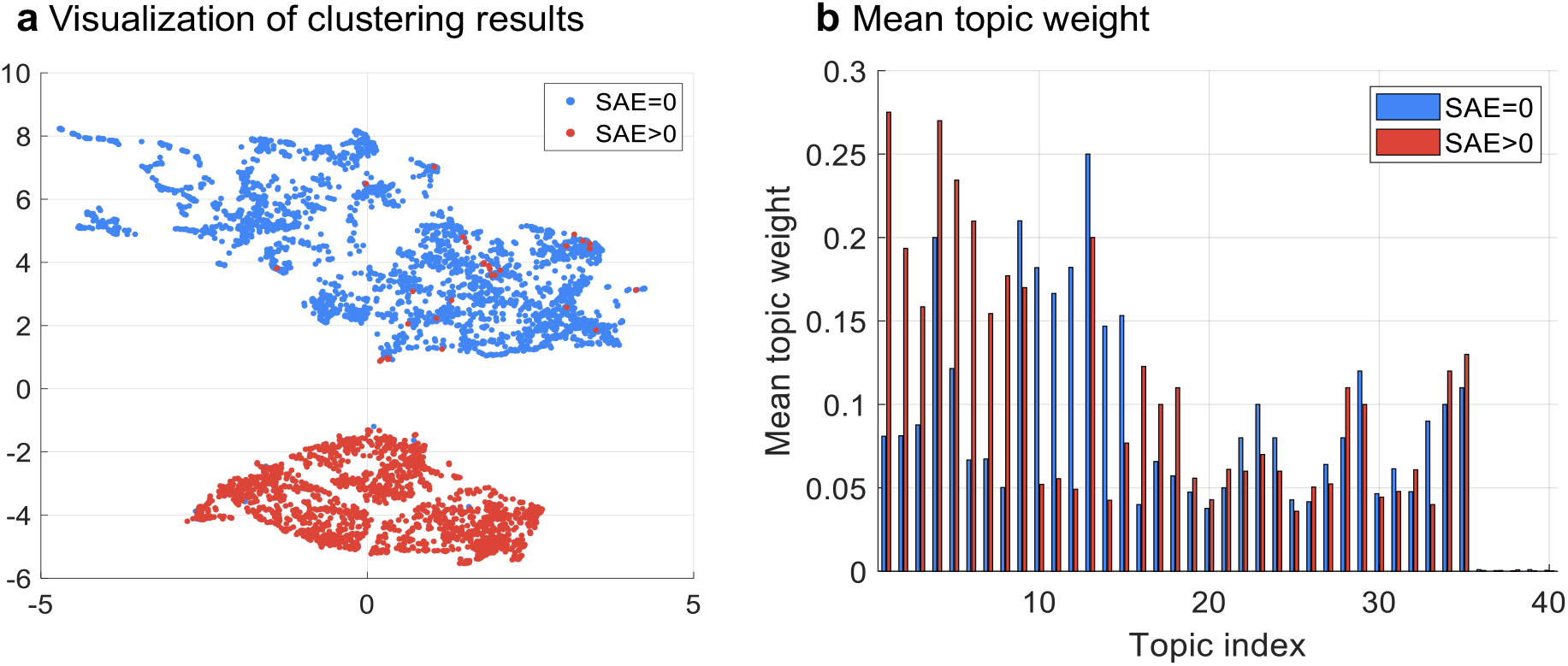
Donepezil clinical trial. **a** Visualization of clustering results; **b** Mean topic weight (MTW) of all topics on two groups, where x-axis is the topic index and y-axis is the MTW of each topic on two subgroups.

We examined the distribution of the 40 topics across the two subgroups (Fig 4b). According to i) the mean topic weights (MTW) over all samples (evaluation for the usage of each topic on the cohort) and ii) Mann–Whitney U (MWU) test (18) on MTW of SAE subgroup (#SAE>0) versus non-SAE subgroup (#SAE=0) (evaluation for difference of topic usage between two groups), 18 typical topics were selected to be analyzed detailly. Specifically, 15 topics (except for T4, T9, and T13 in Fig 2c) whose MWU p-value of SAE subgroup versus non-SAE subgroup are smaller than 0.05 (significant difference between two groups) while MTW on all samples are larger than 0.1 were selected. Of the 15 typical significant-difference topics, 10 topics (T1∼T3, T5∼T8, T16∼T18, denoted by red in Fig. 3c) were characterized as likely to align with the SAE subgroup (#SAE>0) and the other 5 topics (T10∼T12, T14∼T15, denoted by blue in Fig. 3c) align with the other non-SAE subgroup (#SAE=0). Besides, 3 topics (T4, T9, and T13) whose p-values are larger than 0.05 but MTW on all samples are the top-three largest ones were also selected. These 3 topics (T4, T9, and T13) are shared by all clusters.

We then examined the relevance of our learned disease topics by qualitatively assessing the coherence of the five most prevalent clinical events (i.e., diagnosis and medication codes) for each topic (19). We interpreted these 18 topics based on these clinical events and found that many of them were specific to different diseases (Fig 3c). These 18 topics can be divided into three categories. Specifically, i) T4, T9, and T13 includes dementia, memory loss and cognitive impairment related events, which are commonly used among all clusters. These topics represent the common diseases and drugs in the cohort. ii) T1 is related to cardiovascular diseases. T2 is related to gastrointestinal diseases. T3 is about respiratory disorders. T5 is related to psychotic disorders, especially Schizophrenia and relevant treatments (anticholinergic agents) (20). T6 is related to endocrine disorders. T7 is about metabolism disorders such as mineral metabolism disorder. T8 includes lipoid metabolism and secondary malignant neoplasm or cancer of the liver. There have been prior studies showing the relationship between these two types of diseases (21). T16 includes various conditions or disorders of brains, which is closely related to AD. T17 talks about the diseases and treatments of esophagus such as gastroesophageal reflux disease (GERD). T18 is about obesity and some related complications and drugs. Six clusters were derived which is characterized by some related clinical topics: cluster 1 (N=1811; 36.23%), patients with disorders of ears or eyes (T11 and T14); cluster 2 (N=939; 18.79%), patients with diseases of urinary system (T12 and T15); cluster 3 (N=331; 6.62%), patients with depression or mood disorder (T10 and T13); cluster 4 (N=667; 13.35%), patients with disorders of endocrine and metabolism (T6, T7, and T8); cluster 5 (N=548; 10.96%), patients with different diseases of brain (T1, T5, and T16); cluster 6 (N=702; 14.05%), patients with diseases of digestive and respiratory systems (T2, T3, and T17).

The topics, commonly used in the SAE subgroup, may have close relation with exclusion criteria in EC. T11 and T14 are related to diseases of ears such as hearing loss in T11 and otitis media in T14 with related drugs, which have some shared characteristics with AD (22). T12 and T15 related to various diseases are about male genital organs such as hyperplasia of prostate and urinary tract disorder. T10 is associated with mood disorders and depression, where their relationship to dementia and AD is unclear (23). These topics are mostly related to relatively mild conditions with no direct connection to the diagnosis of AD. In other words, most phenotypes associated with the non-SAE subgroup are mild aging-related chronic comorbidities that are common in older adults and do not lead to serious adverse effects. Therefore, diseases which are more associated with the non-SAE patient subgroup may have a lower probability of causing adverse serious events.

We further analyzed the association between the inferred topics with the SAE subgroup and the extracted computable eligibility criteria. We will explain why our model is more powerful in distinguishing two subgroups compared to traits.

### Align with exclusion criterion in clinical trials

Firstly, we found that topics (T1∼T8) aligned with the SAE patient subgroup (#SAE>0) are highly associated with the exclusion criteria of the donepezil trial. Here, [Ex] represents the “exclusion criterion” of clinical trials.

- *“[Ex] Patients with evidence of clinically significant, active gastrointestinal, renal, hepatic, respiratory, endocrine, or cardiovascular system disease (including history of life-threatening arrhythmias)*.*”* We found T1 (cardiovascular), T2 (gastrointestinal), T3 (respiratory), T6 (endocrine) are highly related to this exclusion criterion. Disorders of lipoid metabolism (i.e., T8) is a major contributing factor that mediates the development of cardiovascular diseases, especially atherosclerosis (24).
- *“[Ex] Patients with dementia complicated by other organic diseases or Alzheimer’s disease with delirium*.*”* We found T4 (delirium) is related to this criterion. Although T4 is a topic shared by non-SAE subgroup (cluster 1) and SAE subgroup (clusters 4, 5, 6), its proportion and MTW are higher in SAE group (Fig 3b and 4b).
- *“[Ex] Patients with psychiatric disorders affecting the ability to assess cognition such as schizophrenia, bipolar or unipolar depression. Patients with clinically significant sleep disorders will also be excluded unless these are controlled by treatment and clinically stable for > 3 months prior to screening*.*”* We found T5 (psychotic disorders) is related to this criterion.
- *“[Ex] Patients with any conditions affecting absorption, distribution, or metabolism of the study medication (e*.*g*., *inflammatory bowel disease, gastric or duodenal ulcers, hepatic disease, or severe lactose intolerance)*.*”* We found T7 is related to this criterion. T7 is characterized as other disorders of metabolism include disorders of fluid, electrolyte, and acid-base balance. Acid-base and electrolyte homeostasis is essential for the proper functioning of many metabolic processes and organ functions in the body (25). Phenotypes in T7 seem to have effects on absorption or metabolism of the study medication.
- *“[Ex] Patients with a history of cancer (does not include basal or squamous cell carcinoma of the skin) treated within 5 years prior to study entry, or current evidence of malignant neoplasm, recurrent, metastatic disease. Males with localized prostate cancer requiring no treatment would not be excluded*.*”* Although we annotated T8 to the disorder of lipoid metabolism, it also includes secondary malignant neoplasm or cancer of the liver. As the disorder of lipid metabolism plays an important role in carcinogenesis and development, we concluded T8 is also related to this criterion.

### Different from and move beyond exclusion criteria in clinical trials

Secondly, we found that some topics (T16∼T18) have no direct connections with exclusion criteria. These topics describe some diseases, such as disorders of the brain in T16, diseases of esophagus in T17, and obesity in T18, which may have a severe impact for older people. For example, in (26), authors discussed associations of obesity with numerous life-threatening diseases. In (27), authors discussed the potential life-threatening gastroesophageal reflux disease (GERD). In other words, our method may discover some diseases that have potential outcomes of serious adverse events for elder patients, but are ignored by the original EC. This is the first reason why our method, learning knowledge from more abundant features, can separate SAE and non-SAE subgroups well compared with original traits from EC. Furthermore, even for one disease that appears in both SAE-associated topics and EC, more abundant features provide more detailed insights. For example, for gastrointestinal disease, EC only said “[Ex] *Patients with evidence of clinically significant active gastrointestinal disease”*, which may be a coarse description. However, the learned topics, T2 and T17, discover more detailed diseases or drugs about gastrointestinal disease.

There are also eligibility criteria that have no apparent relations with these inferred topics likely due to two main reasons: (1) since we did not use procedure codes and laboratory tests in the topic modeling process, eligibility criteria related to procedures and lab tests were missing in these inferred topics; and (2) there are eligibility criteria that are not used to ensure patient safety but to ensure treatment efficacy.

### Target population of the bevacizumab (Avastin) clinical trial

To demonstrate the generalizability of our approach, we analyzed another RCT (NCT00112918) which studied a combination chemotherapy with or without bevacizumab in treating patients who have undergone surgery for high-risk stage II or stage III colon cancer. Specifically, the trial studied three combination chemotherapies: (1) the control arm, a standard-of-care (SoC) treatment, FOLFOX4 (i.e., intermittent fluorouracil/leucovorin with oxaliplatin), (2) FOLFOX4 plus bevacizumab, and (3) XELOX (i.e., capecitabine with oxaliplatin) plus bevacizumab. In our RWD analysis, we set the target population as patients who (1) were diagnosed with colorectal cancer (CRC), and (2) treated with FOLFOX4 (Fig. 5(b), Table 2). Since FOLFOX4 was the SoC treatment, patients who were treated with FOLFOX4 in the real-world could potentially benefit from the new combination chemotherapies investigated in this trial. A total of 739 patients (mean age, 57.49 with a standard deviation of 11.2 years old) were identified (out of a total of 47,492 CRC patients) from OneFlorida.

**Table 2.** Demographic characteristics and selected traits of the target population of the bevacizumab (Avastin) clinical trial for colorectal cancer.

| Characteristic | Overall<br>(N=739) | # SAEs = 0<br>(N=347) | # SAEs > 0<br>(N=392) | $\chi^2$<br>p-value | Adjust Demo* |
| --- | --- | --- | --- | --- | --- |
| **Age**, Mean (SD), yr | 57.49 (11.2) | 59.13 (11.2) | 56.0 (11.1) |  |  |
| **Sex**, No. (%) |  |  |  |  |  |
| Female | 328 (44.3) | 141 (40.6) | 187 (47.7) |  |  |
| **Race/Ethnicity**, No. (%) |  |  |  |  |  |
| White | 488 (66.0) | 237 (68.3) | 251 (64) |  |  |
| Black | 172 (23.3) | 79 (22.8) | 93 (23.7) |  |  |
| Asian | 10 (1.4) | 5 (1.4) | 5 (1.2) |  |  |
| Others & Unknown | 69 (9.3) | 26 (7.5) | 43 (10.9) |  |  |
| **traits**, No. (%) |  |  |  |  |  |
| Colon carcinoma | 616 (83.3) | 296 (85.3) | 320 (81.6) | 0.182 | 0.058 |
| Metastatic disease | 499 (67.5) | 221 (63.6) | 278 (70.9) | 0.036 | 0.026 |
| parenteral anticoagulants | 240 (32.4) | 77 (22.1) | 163 (41.5) | $\leq 0.001$ | $\leq 0.001$ |
| immunotherapy | 146 (19.7) | 75 (21.6) | 71 (18.1) | 0.233 | 0.130 |
| anti-angiogenic treatment | 137 (18.5) | 72 (20.7) | 65 (16.5) | 0.146 | 0.081 |
| myocardial infarction | 90 (12.1) | 25 (7.2) | 65 (16.5) | $\leq 0.001$ | $\leq 0.001$ |
| Significant traumatic injury | 40 (5.4) | 16 (4.6) | 24 (6.1) | 0.365 | 0.379 |
| thrombolytic agent | 38 (5.1) | 9 (2.5) | 29 (7.3) | 0.003 | 0.003 |
| central nervous disease | 37 (5) | 12 (3.4) | 25 (6.3) | 0.070 | 0.099 |
| inability to take oral medication | 33 (4.4) | 10 (2.8) | 23 (5.8) | 0.050 | 0.034 |
| Open biopsy | 30 (4) | 17 (4.8) | 13 (3.3) | 0.277 | 0.282 |
| radiotherapy | 24 (3.2) | 10 (2.8) | 14 (3.5) | 0.598 | 0.790 |
| bone fracture | 21 (2.8) | 8 (2.3) | 13 (3.3) | 0.410 | 0.392 |
| coagulopathy | 20 (2.7) | 8 (2.3) | 12 (3) | 0.528 | 0.672 |
| oophorectomy | 17 (2.3) | 7 (2) | 10 (2.5) | 0.630 | 0.956 |
| cerebrovascular accidents | 14 (1.8) | 5 (1.4) | 9 (2.2) | 0.396 | 0.428 |

**Fig. 5.**
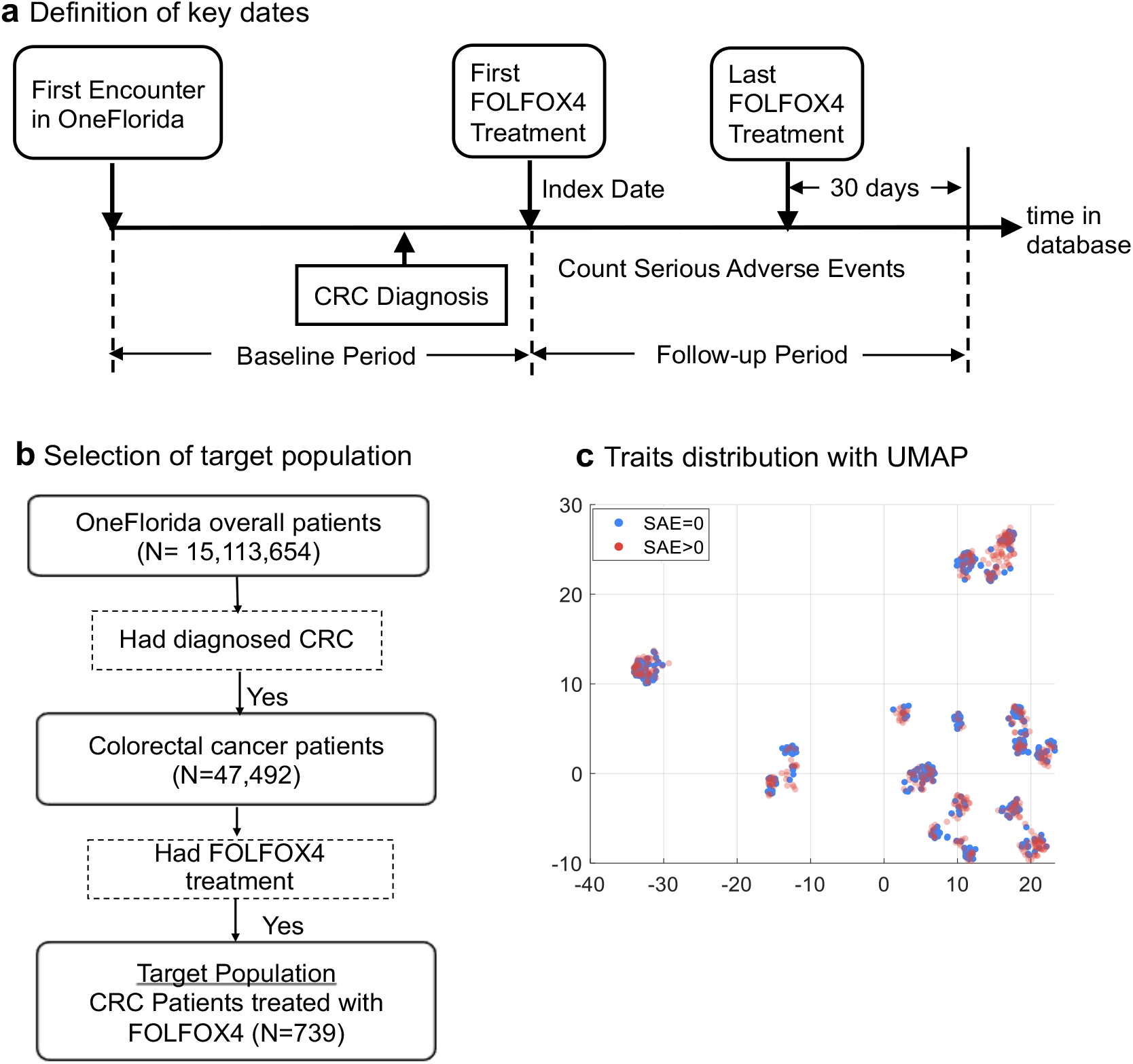
The Bevacizumab (Avastin) clinical trial for colon cancer. **a** Definition of key dates; **b** Selection of the target population; **c** Traits distribution with UMAP among the two patients’ groups with or without SAEs.

Similar to the Donepezil trial, we defined the index date as the date of first FOLFOX4 treatment after CRC diagnosis. The baseline period was defined as the time period before the index date, and the follow-up period was defined as the time period between index date and the last FOLFOX4 treatment date plus 30 days. SAE events were collected from the follow-up period. This study design is depicted in Fig. 4(a). Among all CRC patients who received FOLFOX4 after diagnosis, 347 (47.0%) had no SAE, while 392 (53.0%) had at least one SAE. We examined the computability of each eligibility criterion and constructed queries to extract study traits corresponding to the computable ones based on the OneFlorida data. The identified traits included patient demographics (e.g., age) and medical history (e.g., medication, treatment). We conducted Chi-square tests on the two patient subgroups, i.e., patients had SAEs (#SAE>0) vs patients who did not (#SAE>0). We found that the p values of most study traits are larger than 0.05, except for metastatic disease (p=0.026), parenteral anticoagulants (p<0.001), myocardial infarction (p<0.001) and significant and thrombolytic agent (p=0.003). The p value of the trait related to “inability to take oral medication” was smaller than 0.05 after adjusting for demographics (Table 2).

Similar to the donepezil trial, we applied SPFA to the CRC target population. The patient features we collected from the baseline period include demographics, diagnosis (as phecodes) and medications (as ATC classes)). Similar to what we have done in analysis of Donepezil trail, we evaluated the topic coherence by NPMI value (15), and the classification performance by ROC-AUC to decide the number of topics. We set the number of topics K=40 since it achieves higher ROC-AUC and NPMI (Supplementary Fig. 1). Fig. 6a shows the UMAP embeddings of new patient representations induced by SPFA. From the figure we can observe two well-separated patient subgroups, which can be identified by K-means clustering. One group (red) is mostly associated with SAE (317 of 347 patients, or 91.4% of the patients in this subgroup encountered SAE) and the other group (blue) is free of SAE (393 patients). We checked the patient group proportions for 40 topics in the two clusters (Fig. 6b). Of the 40 topics in the model, patients’ proportion in 9 topics were larger than 0.075 and visually very different in two patient subgroups. Of these 9 typical topics, 6 topics (T1∼T6) were characterized as likely to align with the SAE subgroup (#SAE>0) and the other 3 topics (T7∼T9) align with the non-SAE subgroup (#SAE=0). We annotated the 9 topics based on the top EHR codes and found that most of the inferred topics were specific to different diseases or treatments (Fig.7, Supplementary Fig. 3).

**Fig. 6.**
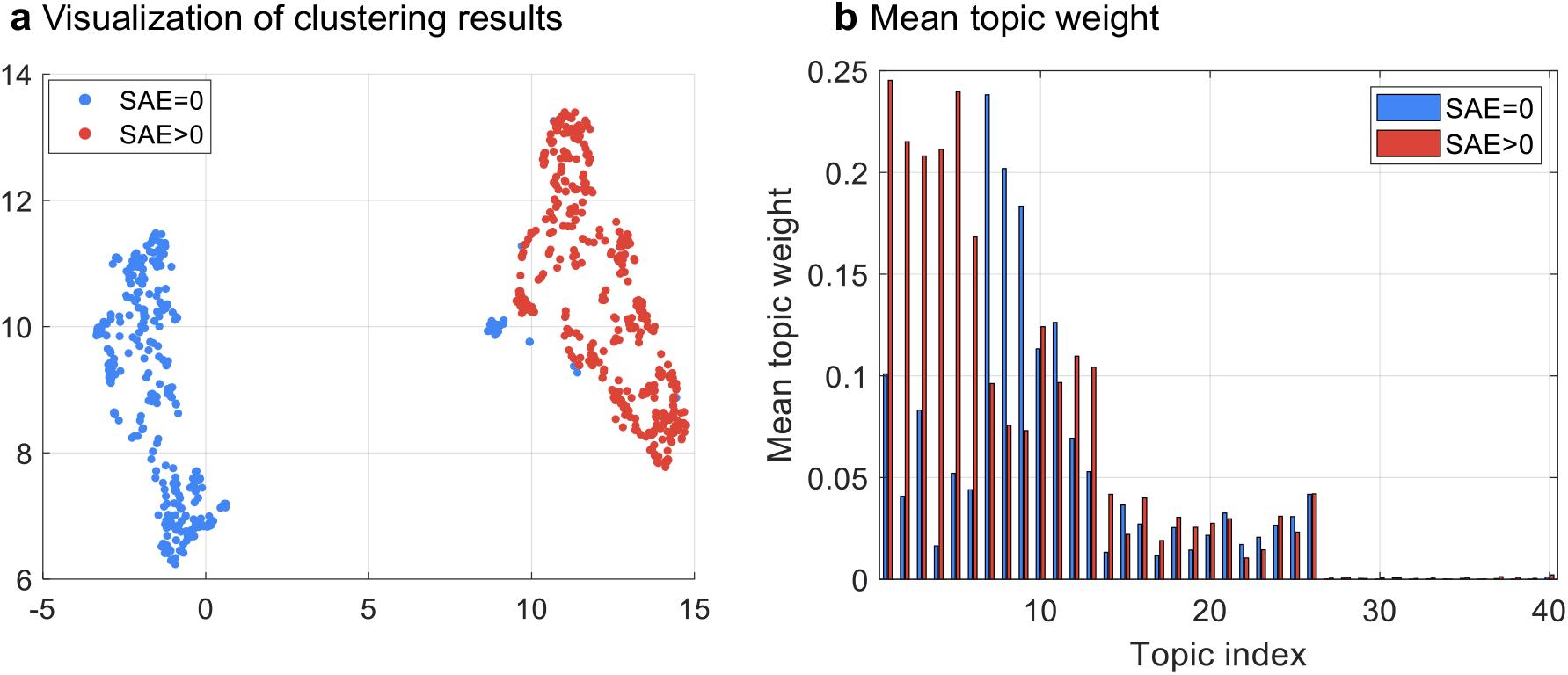
Bevacizumab (Avastin) clinical trial. **a** Visualization of clustering results; **b** Mean topic weight for 40 topics where x-axis is the topic and y-axis is the proportion of each group in that topic.

**Fig. 7.**
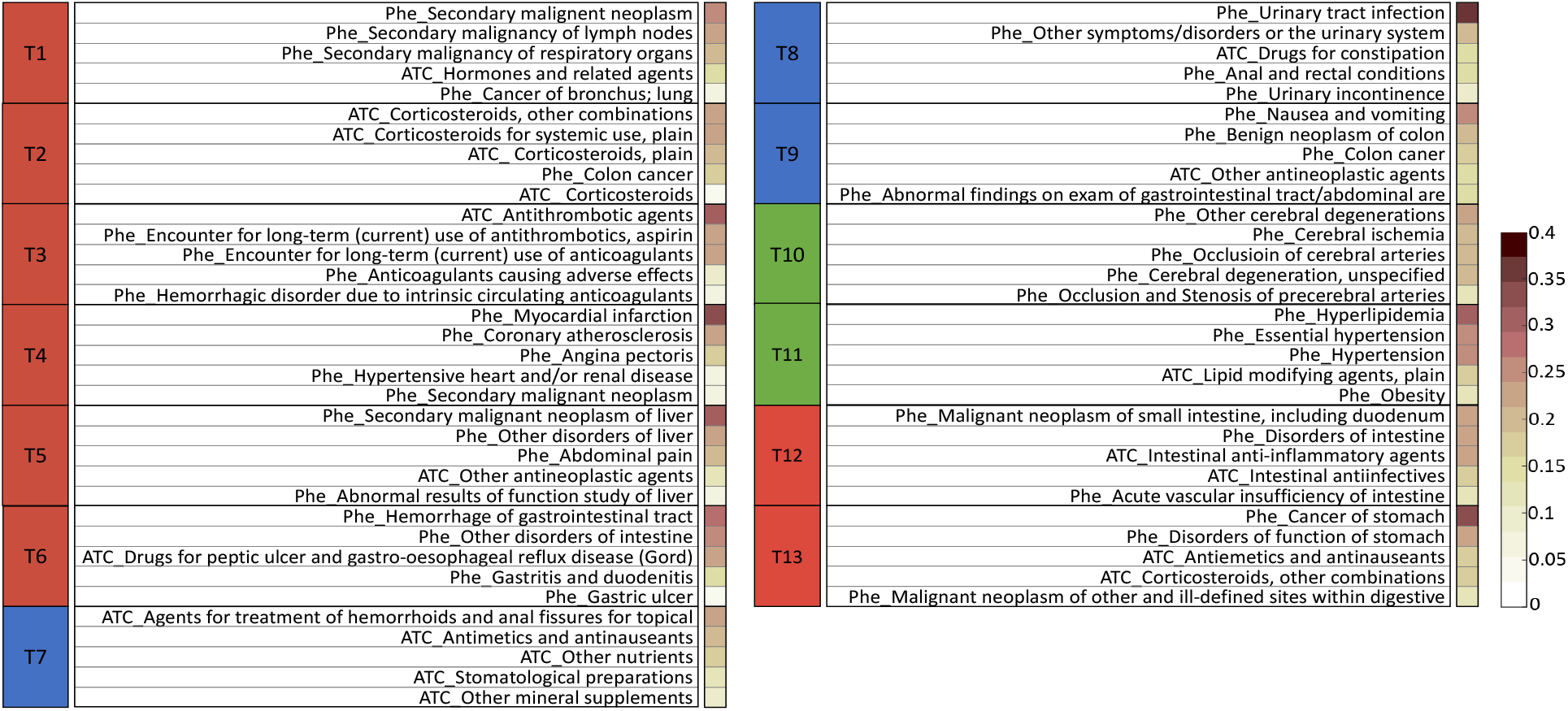
Top clinical features from disease topics in Bevacizumab (Avastin) clinical trial.

Among all 40 topics, similar to the topic selection method for donepezil trial, we selected 13 topics for detailed analysis (Fig. 7). According to the MTW on two groups (Fig. 6b, Supplementary Table 2), these topics can be divided into three categories: i) T1∼T6 and T12∼T13, represented as red, are associated with SAE subgroup; ii) T7∼T9, represented as blue, aligned with the non-SAE subgroup, contain relatively mild diseases and were not directly related to the diagnosis of colon cancer; iii) T10∼T11, represented as green, are often shared on two subgroups. Specifically, T1 is annotated to the use of corticosteroids, with the three of the top five codes being specific corticosteroids treatments. T2 is related to antithrombotic agents. T3 is about malignant neoplasm, where the first three main codes are all correlated with secondary malignant neoplasm and one code is about cancer, and one code is hormones and related preparations drug class which is used to treat cancer. T4 is related to clinically significant (i.e., active) cardiovascular disease. T5 is also related to cancer, but more specific to the liver. Phenotypes in T6 include various kinds of disorders related to intestine, e.g., drugs for peptic ulcer and gastro-oesophageal reflux disease (GORD), hemorrhage of gastrointestinal tract, gastritis and duodenitis and gastric ulcer. T7 includes some commonly used drugs. T8 talks about the disease and treatments about urinary tract infection, which is extremely common in the elderly (28). T9 is associated with gastrointestinal diseases such as nausea and vomiting. T10 and T11 are some common diseases such as or hyperlipidemia or hypertension. T12 includes different diseases or treatments about the intestine. For example, authors in (29) discussed the relations between CRC and chronic intestinal inflammation. T13 is about the cancer or malignant neoplasms of digestive organs. Next, we analyze the connections of SAE-group related topics (the red one) with the EC in trial.

### Association analysis between topics and the eligibility criteria of bevacizumab (Avastin) trial

Firstly, we analyzed the association between the inferred topics and extracted eligibility criteria. We compared each typical topic with the criteria. We found that topics, T1∼T6, aligned with the SAE subgroup (#SAE>0) are highly associated with some of the exclusion criteria in the original trial.

- *“[Ex] Chronic treatment with corticosteroids (dose of* ≥ *10 mg/day methylprednisolone equivalent) (excluding inhaled steroids)*.*”* We found T1 (*corticosteroids*) is highly related to this exclusion criterion.
- *“[Ex] Current or recent (within 10 days prior to study treatment start) use of full-dose oral or parenteral anticoagulants or thrombolytic agents for therapeutic purposes*.*”* We found T2 (antithrombotic) is related to this criterion.
- *“[Ex] Macroscopic or microscopic evidence of remaining tumour. Patients should never have had any evidence of metastatic disease (including presence of tumour cells in the ascites). The isolated finding of cytokeratin positive cells in bone marrow is not considered evidence of metastatic disease for purposes of this study.* *[Ex] Other malignancies within the last 5 years (other than curatively treated basal cell carcinoma of the skin and/or in situ carcinoma of the cervix)*. *[Ex] Previous anti-angiogenic treatment for any malignancy; cytotoxic chemotherapy, radiotherapy or immunotherapy for colon cancer*.*”* We found T3 (malignant neoplasm) and T5 (disorders of liver) are related to these exclusion criteria. Although we annotated T5 as the disorder of liver, it also included malignant neoplasm of liver or antineoplastic agents.
- *“[Ex] Clinically significant (i*.*e*., *active) cardiovascular disease. This includes, but is not limited to, the following examples: cerebrovascular accidents (*≤ *6 months prior to randomization), myocardial infarction (*≤ *1 year prior to randomization)* …*”* We found T4 (cardiovascular) is characterized by myocardial infarction, coronary atherosclerosis, angina pectoris, which is very related to this criterion.
- *“[Ex] Lack of physical integrity of the upper gastro-intestinal tract, malabsorption syndrome, or inability to take oral medication*.*”* We found T6 is related to this criterion.

Secondly, we analyzed the difference of learned topics with EC and discussed why our methods can separate SAE and non-SAE well. Some topics, T12 and T13, are beyond the exclusion criteria in trial, which provide more discriminant features to separate two subgroups. For instance, T12 is about intestinal diseases, which may bring a high mortality rate and often related to multiple comorbidities (30). Even for the topics that have relation with exclusion criteria as discussed above, the learned clinical topics provide more detailed information. For example, EC provides a rough description about corticosteroids as “*Current or recent (within 10 days prior to study treatment start) use of full-dose oral or parenteral anticoagulants or thrombolytic agents for therapeutic purposes”*. Compared with it, the topic T2 contains more detailed drugs about corticosteroids. Therefore, these two benefits make our method provide a better separation for two subgroups.

Similar to the findings in the donepezil trial analysis, there are some criteria which have no relations with the learned topics. Some of these criteria have potential to be adjusted. For example, for the exclusion criterion “*History or evidence upon physical examination of central nervous disease (CNS) disease (e*.*g*., *primary brain tumour, seizure not controlled with standard medical therapy, any brain metastases)*,” Exclusion of patients with brain metastases (BMs) in clinical trials is common. Although life expectancy may be reduced for some patients with BMs, and there are concerns about a potentially greater risk of neurotoxicity, our results did not indicate CRC patients experience a higher rate of SAEs due to having history or evidence upon physical examination of central nervous disease (CNS) disease. If these patients are excluded, justification for such exclusion should be provided alongside the exclusion criteria (31).

## Discussion

Rigorous eligibility criteria for RCTs may make the trial participants not representative of real-world patients. When the trial drugs are on market and prescribed for patients, severe adverse events could happen. In this paper, we developed a machine learning approach to identify the patient subgroups from RWD which are more or less likely to encounter SAEs after the treatment. We consider patient demographics and all clinical events, including diagnosis and medications, in the baseline period for deriving the subgroups. To account for the high-dimensionality and SAE information, we proposed a novel supervised topic modeling approach (SPFA) to achieve this goal. Our approach identified a set of clinical topics and derived novel patient representations based on them, such that the patient subgroups with or without SAEs can be well separated with these representations.

We validated our method on two RCTs from different disease domains: (1) NCT00478205 for AD; and (2) NCT00112918 for colorectal cancer. Tested on both trials, patient subgroup (#SAE=0) and patient subgroup (#SAE>0) can be separated well by k-means clustering using the inferred topics. The inferred topics characterized as likely to align with the patient subgroup (#SAE>0) revealed meaningful combinations of clinical features and can provide data-driven recommendations for refining the exclusion criteria of clinical trials.

In a recent study from Liu et al. (32) which aims at evaluating ECs for oncology trials using RWD and AI, the authors quantified the representability of each study trait with SHAP (33), and they tried to relax the range of each EC for broadening the participation. Only traits with continuous values are considered in a one-by-one manner. Our proposed approach in this paper mainly considered binary traits (continuous traits can also be incorporated with appropriate discretizations followed by one-hot representations) and modeled the high-order interactions of these traits as clinical topics. In addition, we also considered adding extra traits to improve the representability and safety of the trial in real-world data.

Our study has several limitations. First, our study only leveraged the RWD from OneFlorida, which is a regional clinical research network. Future investigation on larger and more diverse RWD is needed to enhance the generalizability of the identified subgroups. Second, we only explored structured information in RWD in this study. Lots of important information, such as symptoms, clinical assessments (e.g., from radiology and pathology reports) and socioeconomic status, are encoded in clinical notes. Extracting and incorporating unstructured information in our study is another important direction to pursue. Third, only discrete traits have been considered in this study. Continuous traits, such as lab tests, are also crucial for many RCTs. Their corresponding computable counterparts in RWD should be explored as well.

## Methods

### Clinical trials

We selected two phase III clinical trials from ClinicalTrials.gov — a registry maintained by the National Library of Medicine (NLM) in the United States.

NCT00478205 is a double-blind, double-dummy trial aims to compare different dosages of donepezil for treating patients with moderate to severe AD (11). We identified AD patients from OneFlorida using ICD-9/10-CM codes (i.e., ICD-9: 331.0; ICD-10: G30.0, G30.1, G30.8, and G30.9). Then, we used the RxNorm concept unique identifier (RXCUI) and National Drug Code (NDC) codes to identify AD patients who took donepezil.

NCT00112918 is a randomized trial studying different combination chemotherapies (i.e. with or without bevacizumab) in treating CRC patients. Our study focused on the control arm (i.e., patients who were treated with the SoC, FOLFOX4). We identified CRC patients using ICD-9/10-CM codes (i.e., ICD-9: 153.*, 154.*, 159.0; ICD-10: C18.*, C19.*, C20.*, C26.0). Within the CRC patients, we then used the RXCUI and NDC codes of three drugs (i.e., 5-fluorouracil, leucovorin, and oxaliplatin) (Supplementary Table 3) to further identify the CRC patients who took FOLFOX4 after their CRC diagnosis.

Real-world patient data (RWD) from OneFlorida: We obtained individual-level patient data from the OneFlorida Clinical Research Consortium (10), which contains robust longitudinal and linked patient-level RWD of ∼15 million (>60%) Floridians, including data from Medicaid claims, cancer registries, vital statistics, and EHRs from its clinical partners. As one of the Clinical Data Research Networks (CDRNs) contributing to the national Patient-Centered Clinical Research Network (PCORnet), OneFlorida includes 12 healthcare organizations that provide care through 4,100 physicians, 914 clinical practices, and 22 hospitals, covering all 67 Florida counties. OneFlorida follows the PCORnet Common Data Model (CDM) including patient demographics, enrollment status, vital signs, conditions, encounters, diagnoses, procedures, prescribing (i.e., provider orders for medications), dispensing (i.e., outpatient pharmacy dispensing), and lab results (8).

We extracted the clinical events from OneFlorida data, which include demographics (i.e., age, sex, race), diagnoses (i.e., ICD 9/10), and drugs (i.e., NDC/RXNorm). We calculated age based on the birth date and first donepezil treatment date for AD patients, or first FOLFOX4 treatment date for CRC patients. Uniform-sized bins were used to discretize the age and one-hot encoding was adopted to encode age, gender and race variables. We mapped diagnosis codes to Phecode which is designed to facilitate phenome-wide association studies (PheWAS) in EHRs (34). Drug codes (i.e., NDC/RXNorm) were mapped to the Anatomical Therapeutic Chemical (ATC) Classification System 3rd level. Finally, we concatenated all the features (demographics, diagnosis and medications) to represent each patient as a binary vector.

### Defining serious adverse events (SAEs)

To define an SAE, we followed the Food and Drug Administration (FDA) Code of Federal Regulations (CFR) Title 21 definition of SAE (35). An adverse event (AE) is considered serious if, in the view of either the investigator or sponsor, it results in any of the following outcomes: 1) death; 2) a life-threatening adverse event, 3) inpatient hospitalization or prolongation of existing hospitalization, 4) disability or permanent damage, 5) congenital anomaly/birth defect and 6) important medical events (IME) that may not result in death, be life threatening, or require hospitalization may be considered a serious adverse drug experience when, based upon medical judgment, they may jeopardize the patient or subject and may require medical or surgical intervention to prevent one of the outcomes listed in this definition. Another resource for defining SAE is the Common Terminology Criteria for Adverse Events (CTCAE) - a descriptive terminology for AE reporting (36). CTCAE categorizes AE into Grade 1 (mild), Grade 2 (moderate), Grade 3 (severe or medically significant but not immediately life-threatening), Grade 4 (life-threatening consequences), and Grade 5 (death). Considering both the definition from FDA CFR title 21 and CTCAE, we defined an AE as SAE if it results in hospitalization or death.

In this study, to identify SAEs for patients treated with Donepezil or FOLFOX4, we first identified the reported SAEs in the Result section of the selected trials from ClinicalTrails.gov. For each SAE, we first collected the ICD-9/10-CM codes to identify corresponding health conditions (Supplementary Table). To further identify SAEs, we mapped these terms to the CTCAE terms and categorized them as SAEs based on the grading scale above (i.e., CTCAE Grade 3/4 and 5).

### Poisson factor analysis

By collecting all patient vectors, we can construct a binary data matrix *X* ∈ {0, 1}^*V* ×*N*^, with *V* corresponding to the number of features and *N* being the number of patients. Poisson factor analysis (PFA) (37) assumes *X* following a Poisson likelihood as

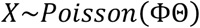

where 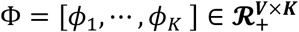 is the topic matrix with each column *ϕ*_*k*_ being the *k*-th clinical topic, and *ϕ*_*k*_ is a distribution over features; 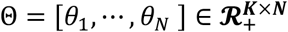 is the topic weight matrix and each column *θ*_*n*_ represents the topic weights of the *n*-th patient. Based on the expectation rule, we have the equation:

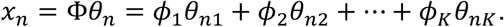

Clearly, each patient vector is composed of weighted summation of all topics, where values in *θ*_*n*_ denotes the weights. Therefore, we call *θ*_*n*_ as topic weights, a new representation for *x*_*n*_, since it exhibits the weight (or proportion after normalization) of each topic in representing the patient *x*_*n*_. We will perform clustering on the learned new representations.

Compared with latent Dirichlet allocation (LDA) (38), which models the distribution of topic weights as Dirichlet distribution, PFA models it as Gamma distribution. The advantage of Gamma distribution for topic weight is that it introduces the shrinkage mechanism to prune inactive factors and enhances the model interpretability (39). From Fig. 4b and Fig. 6b we can see that although we set the number of topics as 40 for both cases, after learning, our model automatically truncates it to 35 for AD and 26 for CRC. It is in accordance with the fact that CRC has less samples which thus can be described with less topics.

### Clustering with supervised PFA models

The original PFA is purely unsupervised. In order to incorporate the outcome information (i.e., having SAE or no) into the topic learning process, we extended the original PFA model to a supervised setting which uses the occurrence of SAE as the supervision information to guide the learning process of PFA. Specifically, for the *n* -th patient, if he/she did not encounter any SAE in the follow-up period, we set *y*_*n*_ = 0; otherwise, we set *y*_*n*_ = 1. Then we adopted the mean-field variational Bayes method (40) to maximize the evidence lower bound (ELBO) of the data likelihood as

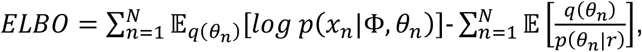

where *p*(*x*_*n*_|Φ, *θ*_*n*_) and *p*(*θ*_*n*_|r) are the Poisson likelihood and Gamma prior as in PFA, *q*(*θ*_*n*_) is the variational posterior to be learned. Currently, we built *q*(*θ*_*n*_) as an encoder network *q*_*W*_(*θ*_*n*_|*x*_*n*_), where *W* represents learnable parameters of the encoder network, and *q*(·) is modeled as a Weibull distribution that makes *θ*_*n*_ positive and sparse (41).

To perform supervised learning, we added a supervised regularizer in the original ELBO objective as

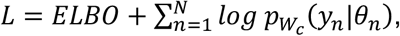

where the second term can be viewed as the label likelihood implemented by cross entropy loss. The model structure of supervised PFA is shown in Figure 1. As a result, we built a probabilistic auto-encoding supervised topic model, whose parameters were encoder parameters *W*, decoder parameters Φ (topics), and classifier *W*_*c*_. We deployed stochastic gradient descent to learn *W* and *W*_*c*_, and stochastic gradient based Monte Carlo Markov Chain sampling to infer Φ (41). Our proposed model can be learned in a mini-batch style, which is easily amenable for large-scale data analysis.

### Mean topic weight (MTW) and Mann–Whitney U (MWU) test

In our analysis, we use MTW to select typical topics and plot the topic distributions on each cluster (pie plot in Fig. 3b). According to the data generation process of PFA and equation (1), topic weight of *n*-th patient *θ*_*n*_ represents the weights of all topics in representing one patient. For fair evaluation, we normalize *θ*_*n*_ as 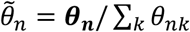 to a Dirichelt distribution (38). As a result, 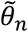 can be regarded as topic proportions. Given a group with *N*′ patients, the MTW of *k*-th topic within this group is calculated as 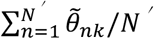. For each topic, after calculation of MTW on SAE subgroup and non-SAE subgroup, we use MWU test (18) to calculate the p-value of each topic for evaluating the significant difference of topic weights on two subgroups.

## Data Availability

The real data analyzed in this article were provided by OneFlorida Clinical Research Consortium and restrictions apply to the availability of these data. Requests for access to the data should be submitted to and approved by OneFlorida Clinical Research Consortium.

## Code availability

The implementation of the proposed algorithm in python are publicly available from https://github.com/haozhangWCM/Submission-to-NC-SPFA-for-EC.

## IRB Information

This study has been reviewed by the University of Florida Institute Review Board under protocols IRB202003137 and IRB202000704.

## Acknowledgements

The work from JX, HZ and FW was supported by NSF 1750326, ONR N00014-18-1-2585 and NIH RF1AG072449. The work from JB and HZ was supported by NIH R21AG068717 and NIH R21CA253394.

## Author contributions

FW and BJ conceived the study. JX and HZ designed and implemented the algorithm. HZ analyzed the data. JX and HZ drafted the manuscript. All authors polished and proofread the manuscript.

