## Supplementary for "Machine learning enabled subgroup analysis with real-world data to inform better clinical trial design"

**^1^Department of Population Health Sciences, Weill Cornell Medicine, New York, NY, USA**

**^2^Department of Health Outcomes and Biomedical Informatics, College of Medicine, University of Florida, Gainesville, FL, USA**

### Supplementary Figures

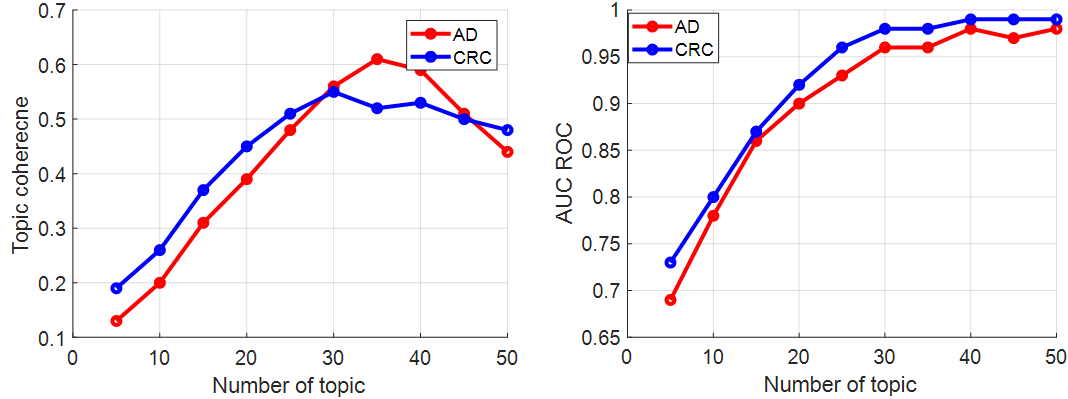

#### Figure 1: The variations of topic coherence and classified AUC-ROC with the number of topics on AD and CRC, which help us to decide the number of topics. With the appropriate number of topics, we hope the model can achieve largest AUC-ROC (separate SAE subgroup (#SAE>0) and non-SAE subgroup (#SAE=0) well) with large topic coherence (good quality of learned topics). Therefore, we choose the number of topics as 40 for both datasets.

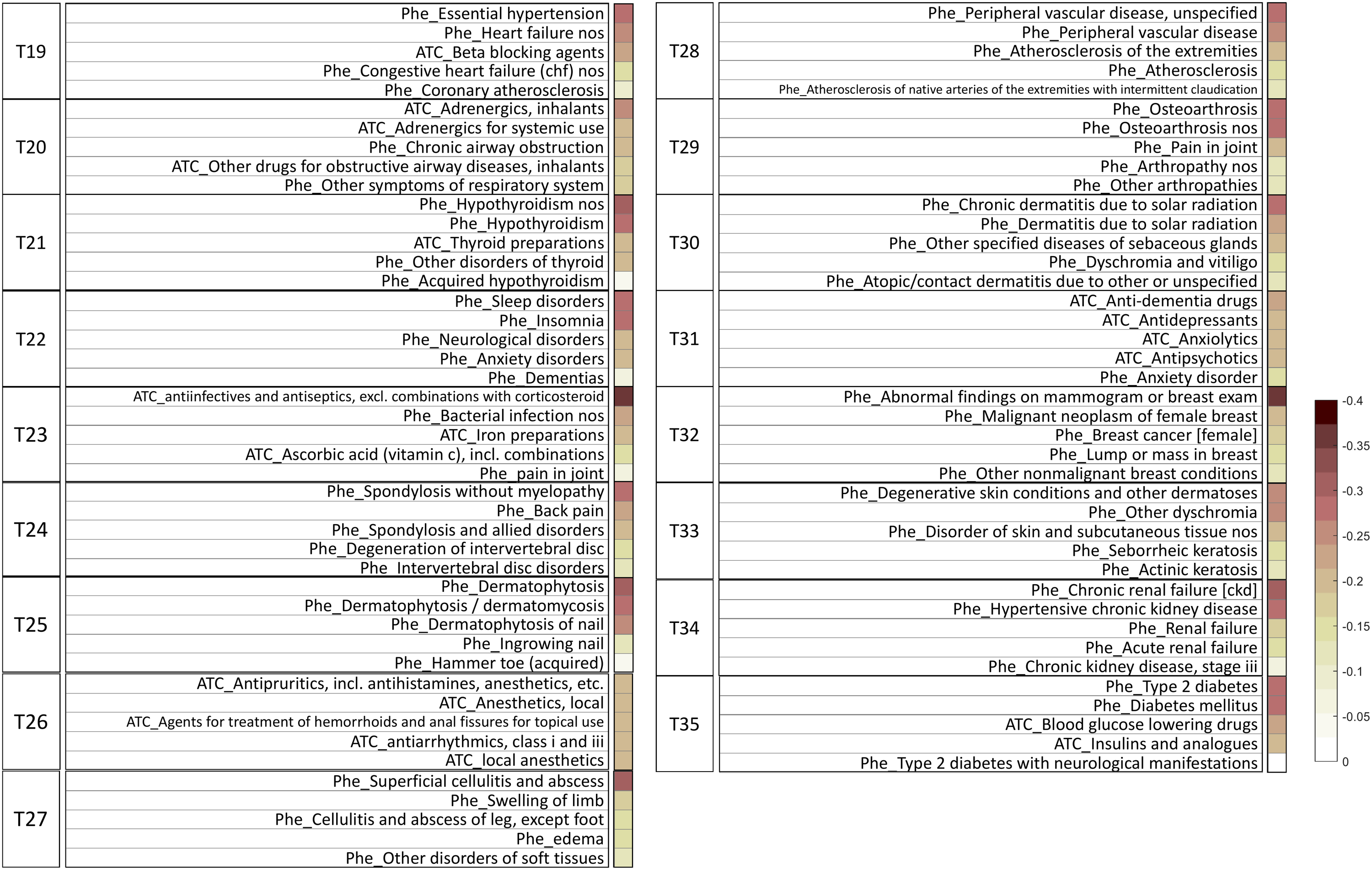

#### Figure 2: Top clinical features from certain disease topics (AD), which is the supplement for Figure 3 in the main manuscript.

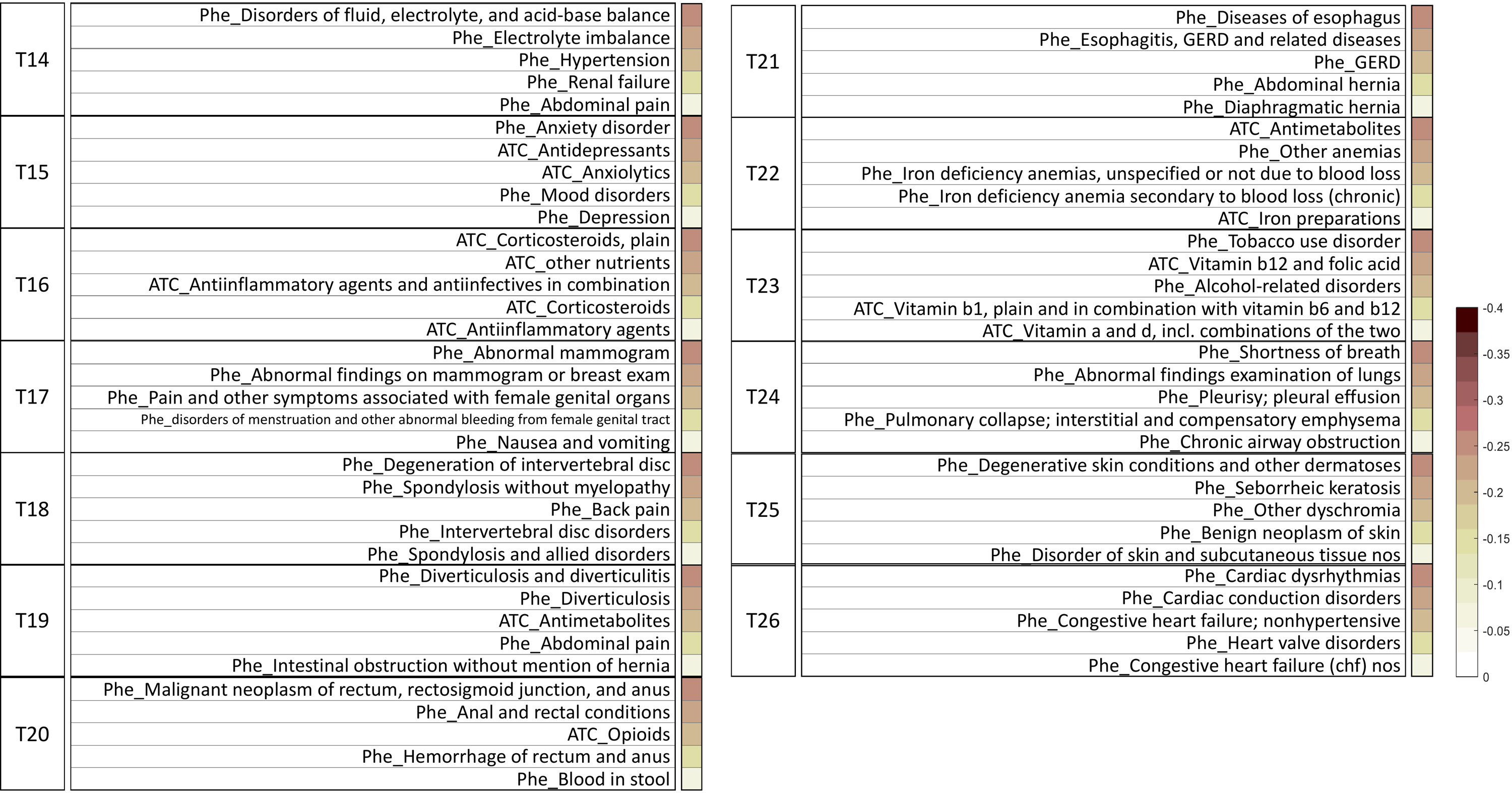

#### Figure 3: Top clinical features from certain disease topics (CRC), which is the supplement for Figure 7 in the main manuscript.

### Supplementary Tables

#### Table 1: On AD dataset, for each topic, we calculate the Mean topic weight (MTW) over all samples, and p-value of Mann–Whitney U test between SAE subgroup (#SAE>0) and non-SAE subgroup (#SAE=0). We use these two values to choose typical topics.

| **Topic index** | **MTW** | **p-value** | **Topic index** | **MTW** | **p-value** |
| --- | --- | --- | --- | --- | --- |
| T1 | 0.1780 | 0.0041 | T21 | 0.0556 | 0.3568 |
| T2 | 0.1374 | 0.0092 | T22 | 0.0700 | 0.0426 |
| T3 | 0.1231 | 0.0061 | T23 | 0.0850 | 0.0387 |
| T4 | 0.2350 | 0.0753 | T24 | 0.0700 | 0.0197 |
| T5 | 0.1780 | 0.0085 | T25 | 0.0394 | 0.0245 |
| T6 | 0.1383 | 0.0026 | T26 | 0.0461 | 0.0324 |
| T7 | 0.1109 | 0.0015 | T27 | 0.0582 | 0.1387 |
| T8 | 0.1137 | 0.0033 | T28 | 0.0950 | 0.1957 |
| T9 | 0.1900 | 0.1052 | T29 | 0.1100 | 0.1478 |
| T10 | 0.1170 | 0.0089 | T30 | 0.0455 | 0.1754 |
| T11 | 0.1110 | 0.0021 | T31 | 0.0546 | 0.1082 |
| T12 | 0.1156 | 0.0032 | T32 | 0.0543 | 0.2356 |
| T13 | 0.2250 | 0.1657 | T33 | 0.0650 | 0.2903 |
| T14 | 0.1047 | 0.0019 | T34 | 0.1100 | 0.1987 |
| T15 | 0.1150 | 0.0067 | T35 | 0.1200 | 0.2354 |
| T16 | 0.1014 | 0.0074 | T36 | 0.0007 | 0.1453 |
| T17 | 0.1029 | 0.0048 | T37 | 0.0005 | 0.0664 |
| T18 | 0.1016 | 0.0083 | T38 | 0.0006 | 0.0349 |
| T19 | 0.0516 | 0.0264 | T39 | 0.0007 | 0.0279 |
| T20 | 0.0403 | 0.3120 | T40 | 0.0004 | 0.0667 |

#### Table 2: On CRC dataset, for each topic, we calculate the Mean topic weight (MTW) over all samples, and p-value of Mann–Whitney U test between SAE subgroup (#SAE>0) and non-SAE subgroup (#SAE=0). We use these two values to choose typical topics.

| **Topic index** | **MTW** | **p-value** | **Topic index** | **MTW** | **p-value** |
| --- | --- | --- | --- | --- | --- |
| T1 | 0.1731 | 0.0026 | T21 | 0.0298 | 0.2139 |
| T2 | 0.1280 | 0.0038 | T22 | 0.0105 | 0.0917 |
| T3 | 0.1457 | 0.0013 | T23 | 0.0145 | 0.4765 |
| T4 | 0.1139 | 0.0006 | T24 | 0.0310 | 0.3681 |
| T5 | 0.1459 | 0.0020 | T25 | 0.0232 | 0.2573 |
| T6 | 0.1062 | 0.0042 | T26 | 0.0420 | 0.7825 |
| T7 | 0.1672 | 0.0059 | T27 | 0.0005 | 0.0215 |
| T8 | 0.1388 | 0.0119 | T28 | 0.0008 | 0.3176 |
| T9 | 0.1283 | 0.0093 | T29 | 0.0003 | 0.2794 |
| T10 | 0.1187 | 0.2476 | T30 | 0.0006 | 0.3071 |
| T11 | 0.1115 | 0.4310 | T31 | 0.0007 | 0.0275 |
| T12 | 0.1195 | 0.0037 | T32 | 0.0004 | 0.2871 |
| T13 | 0.1087 | 0.0021 | T33 | 0.0005 | 0.0015 |
| T14 | 0.0275 | 0.0038 | T34 | 0.0001 | 0.2761 |
| T15 | 0.0293 | 0.0632 | T35 | 0.0008 | 0.0983 |
| T16 | 0.0336 | 0. 1702 | T36 | 0.0002 | 0.3781 |
| T17 | 0.0154 | 0.0638 | T37 | 0.0013 | 0.0530 |
| T18 | 0.0280 | 0.0918 | T38 | 0.0010 | 0.2810 |
| T19 | 0.0200 | 0.1276 | T39 | 0.0004 | 0.0432 |
| T20 | 0.0275 | 0.3692 | T40 | 0.0021 | 0.7297 |
